# Prevalence and antimicrobial sensitivity patterns of uropathogens, in Tikur Anbessa specialized hospital emergency department Addis Ababa, Ethiopia

**DOI:** 10.1101/2022.12.15.22283508

**Authors:** Yared Boru, Dominick Shelton, Aklilu Azazh, Hywet Engida, Fitsum Kifle, Finot Debebe

## Abstract

**Background:** Empirical treatment of infections remains a major contributing factor to the emergence of pathogens that are resistant to antibiotics. The study aimed to assess the prevalence and anti-microbial sensitivity patterns of uropathogens in the emergency center of Tikur Anbessa Hospital, Ethiopia.

**Methods:** Urine sample data collected over two years from January 2015 to January 2016 at Tikur Anbessa Hospital’s laboratory were retrospectively analyzed for bacterial pathogens, and their antimicrobial susceptibility. Antimicrobial sensitivity tests were done using the disc diffusion technique as per the standard of the Kirby-Bauer method.

**Result:** Of the total 220 samples that were collected, 50 (22.7%) were culture-positive. Male to female data ratio was 1:1.1. Escherichia coli was the dominant isolate (50%) followed by Enterococcus species (12%), Enterobacter species (12%), and Klebsiella species (8%). Overall resistance rates to Cotrimoxazole, Ampicillin, Augmentin, and Ceftriaxone were 90.4%, 88.8%, 82.5%, and 79.3%, respectively. The sensitivity rates for Chloramphenicol, Amikacin, Vancomycin, Meropenem, Cefoxitin, and Nitrofurantoin ranged from 72% - 100%. The antibiogram of isolates showed that 43 (86%) isolates were resistant to two or more antimicrobials, and 49 (98%) were resistant to at least one antibiotic.

**Conclusion and recommendation:** Urinary tract infections are mostly caused by Gram-negative bacteria predominantly in females and Escherichia coli are the most common isolates. Resistance rates to Cotrimoxazole, Ampicillin, Augmentin, and Ceftriaxone were high. Chloramphenicol, Amikacin, Vancomycin, Meropenem, Cefoxitin, and Nitrofurantoin are considered appropriate antimicrobials for the empirical treatment of urinary tract infections in the emergency department. Yet, using antibiotics indiscriminately for patients with complicated UTIs may increase the resistance rate and also lead to treatment failure, hence the prescriptions should be revised following the culture and sensitivity results

## Background

Urinary tract infections (UTIs) are one of the most common types of bacterial infections in humans, occurring both in the community and in healthcare settings. UTI is a term applied to a variety of clinical conditions ranging from the asymptomatic presence of the bacteria in the urine to severe infection of the kidneys with subsequent sepsis [1].

Females are more prone to UTIs than males, a phenomenon explained by their short urethra and its anatomical proximity to the anal orifice [2, 3]. Uropathogens are simple to identify as they mainly come from colonic flora [4], and this serves as a rationale for the empirical treatment of community acquired UTI (CA-UTI) [2, 5]. Empirical treatment, however, restricts the ability to monitor antibiotic response, and predisposes to resistance among uropathogens that cause UTI. [5, 6) Despite their use as practical methods for efficient resource use, empirical management plans must be regularly updated to account for shifting pathogen susceptibility patterns. [6] This is particularly true for developing nations, where the lack of resources for routine antibiotic sensitivity tests compounds an additional challenge [6].

In recent years, the resistance of uropathogens to previously effective antibiotics has become a global phenomenon [7, 8, 9]. Antimicrobial resistance (AMR) is currently estimated to account for more than 700,000 deaths per year worldwide [10]. If no appropriate measures are taken to halt its progress, it is projected to cost approximately 10 million lives and about US$100 trillion per year by 2050 [10] In contrast to other health issues, AMR is a problem that concerns every country irrespective of its level of income and development. The 2014 WHO report identified Africa and South East Asia as the regions without established AMR surveillance systems [11].

The lack of quality data is challenging, resulting in inadequate treatment guidelines for the local situation. The gap in public health capacity is also an issue given the rapidly evolving resistant mechanism of the pathogens and the emergence of multidrug-resistant bacteria that can only be detected through systematic screening in quality-assured microbiology laboratories [12, 13]. Given the widespread irrational use of antibiotics in Africa, the situation is dire [9, 11] as compared to developed nations.

To design suitable interventions, it is important to understand the current status of AMR and identify rollout gaps. The most recent version of the national Ethiopian treatment guideline recommends using Trimethoprim/Sulfamethoxazole (TMP-SMX) and Norfloxacin for the empirical treatment of CA-UTI in Ethiopia, one of the second-most populous regions of Africa [14]. However, we believe that there might be significant antibiotic resistance to the recommended treatments, and as a result, the likelihood of treatment failure is rising with the continued empirical use of these medications. As such, the study aimed to assess antibiotic resistance to both the suggested treatments and other drugs, as well as to identify patterns of resistance to a wide range of potentially helpful alternatives for the treatment of UTIs in Tikur Anbessa Specialized Hospital, Addis Ababa, Ethiopia.

## METHODS

This study was conducted in Tikur Anbessa Specialized hospital. It is the largest referral and tertiary-level teaching hospital in Ethiopia, located in the heart of Addis Ababa, the capital city [15]. The hospital is administered by Addis Ababa University and is the largest tertiary unit and the oldest teaching hospital in Ethiopia, with over 800 beds and provides teaching for about 300 medical students and 350 residents every year and has more than 800 beds [15]. It offers diagnosis and treatment for approximately 370,000-400,000 patients a year [15]. The hospital’s laboratory is one of the biggest and most advanced award-winning laboratories, providing services like culture and sensitivity, gram stain, blood chemistry, Complete Blood Count (CBC), GeneXpert, and Acid-fast bacillus (AFB) for diagnosis of Tuberculosis (TB). The hospital is also known for its multidisciplinary collaborative research with both logical and international universities.

After receiving ethical approval from Addis Ababa University and a letter of support from the department of emergency medicine and critical care, a retrospective two-year record review was carried out on all UTI patients older than 14 years for whom culture and sensitivity tests were performed in the study. This review covered the period from January 2015 to January 2017. The data was retrieved and recorded in a data extraction sheet from the laboratory logbook, which contains patient age, sex, date of collection, department, isolated bacteria, and an antimicrobial susceptibility profile. The data were collected by nurses trained for this purpose. Data was cleaned, coded, and analyzed using the SPSS software program version 21.0.

## RESULTS

### 1. Sociodemographic Information

From Jan 2015 to Jan 2017 a total of 220 urine samples were collected and analyzed. The age of the patients ranged from 14 to 100 years, with a mean age of 32.26 (SD=14.45) years. The mean ages of male and female patients were 35.1 (SD=15.8) and 30.78 (SD=12.4) years, respectively. One hundred nineteen (54%) urine samples were from females and 101(46%) were from male patients, with male to female ratio of 1:1.17 (Table1).

**Table1.** Age and sex distribution of patients with urine culture and sensitivity, Tikur Anbessa specialized hospital emergency department from Jan1 2015 to Dec 30 2016, Addis Ababa Ethiopia

| Sociodemographic Characteristics | Positive no. % | No significant growth % | Negative no. % | Total no. % | P value |
| --- | --- | --- | --- | --- | --- |
| Age (years) 14-25 | 17 (34%) | 11(36.7%) | 47 (33.5%) | 75 (34%) | P=0.34 |
| 26-35 | 8 (16%) | 13(43.3%) | 36 (25.7%) | 57(25.9%) |  |
| 36-45 | 9 (18%) | 2 (6.7%) | 23 (16.4%) | 34(15.4%) |  |
| 46-65 | 15 (30%) | 4 (13.3%) | 34 (24.2%) | 53(24 %) |  |
| >65 | 1 (2%) | 0 | 0 | 1(0.45%) |  |
| Gender Male | 19 (38%) | 8 (26.6%) | 74 (52.8%) | 101 (46%) | P=0.363 |
| Female | 31 (62%) | 22(73.3%) | 66 (47.1%) | 119 (54%) |  |
| Total | 50 (100%) | 30(100%) | 140 (100%) | 220(100%) |  |

### 2. Prevalence of uropathogens

The overall prevalence of uropathogens was 22.7%. The majority of the pathogens (14%) were isolated from females. Most of the uropathogens were isolated from patients in the age group of 14 to 25 years (Table1). E. coli was the most predominant pathogen (50%) isolated from urine samples. (Figure1). The Gram-negative and Gram-positive bacteria were responsible for 84% and 16% of the isolates, respectively.

**Figure1.**
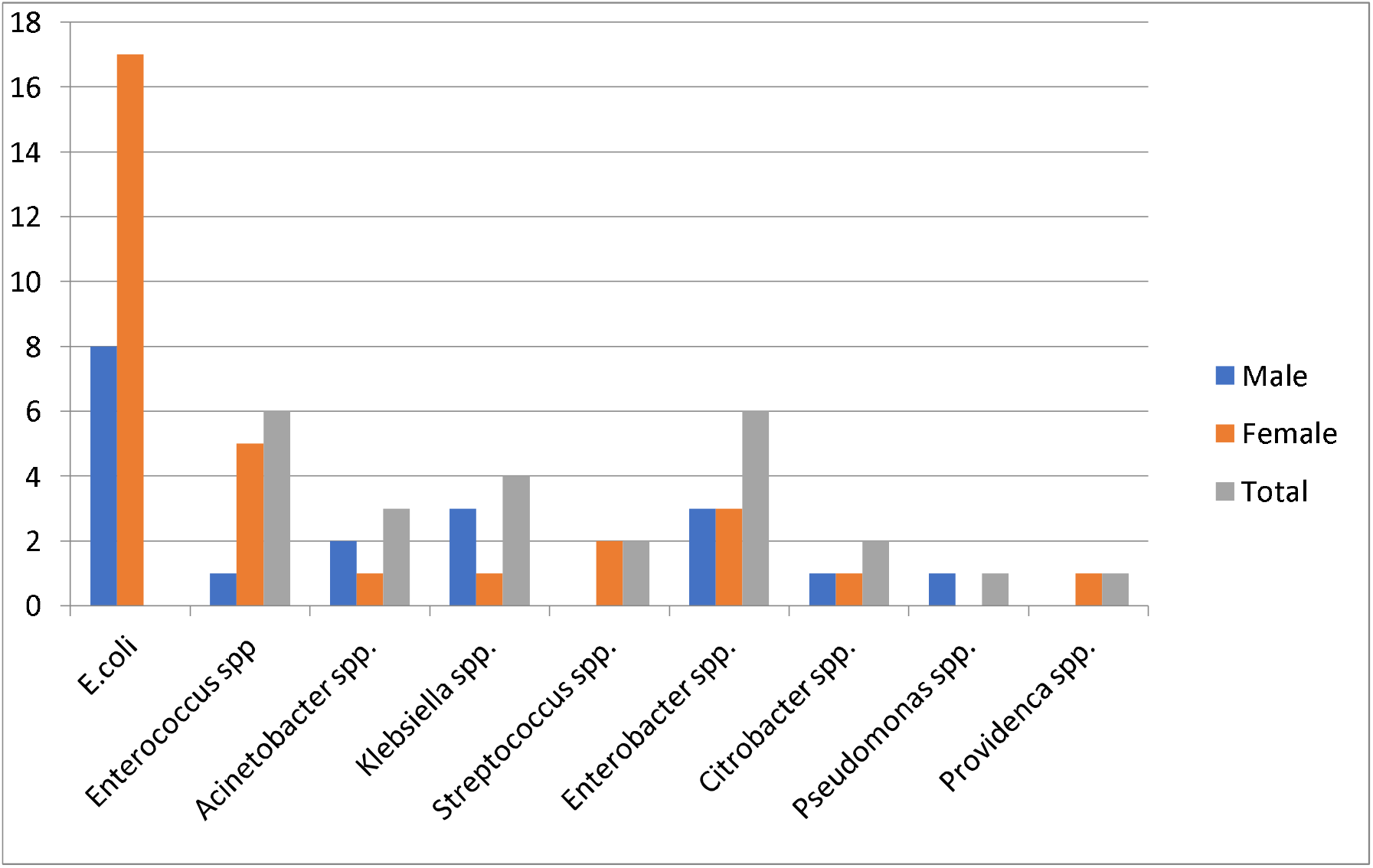
Bacterial isolates of patients who have urine culture and sensitivity done, Tikur Anbessa specialized hospital emergency department from jan1 2015 to Dec 30 2016

### 3. Antibiotic susceptibility pattern of uropathogens

The overall antibiotic susceptibility profiles of bacterial isolates are shown in Table3. Cotrimoxazole had the highest overall resistance (90.4%), followed by Ampicillin (88.8%), Augmentin (82.5%), and Ceftriaxone (79.3%). Meropenem, Amikacin, Vancomycin, and Cefoxitin were all sensitive (Table 2). Because of the non-sustained availability of those antibiotics for which all tasted etiologies are sensitive, only a few organisms were tested. Klebsiella species are dominant in male patients and almost resistant to all antibiotics which shows the risk of having a resistant strain is higher in male patients. Species-specific antimicrobial resistance rates are displayed in (Table3). E.Coli showed high resistance rates to Cotrimoxazole (85.7%), Ampicillin (76.9), and Augmentin (71.4%). The other three most common isolates exhibited resistance patterns of (80% - 100%) to Ampicillin, Cotrimoxazole, and Augmentin.

**Table2.**
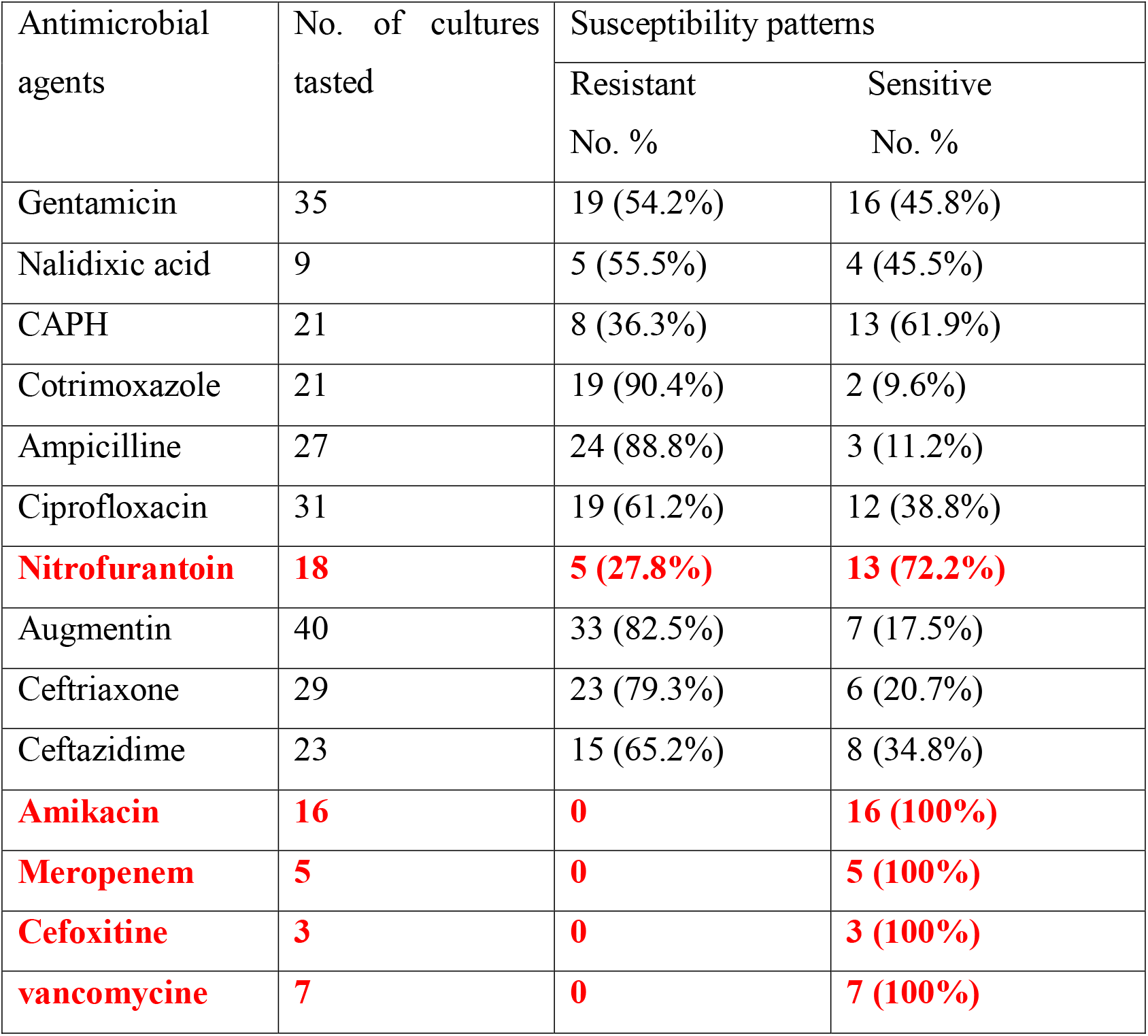
Overall antimicrobial susceptibility profiles of urine samples from patients who have urine culture and sensitivity done, Tikur Anbessa specialized hospital emergency department from, Jan1 2015 to Dec 30 2016, Addis Ababa Ethiopia

| Antimicrobial agents | No. of cultures tasted | Susceptibility patterns |  |
| --- | --- | --- | --- |
|  |  | Resistant<br>No. % | Sensitive<br>No. % |
| Gentamicin | 35 | 19 (54.2%) | 16 (45.8%) |
| Nalidixic acid | 9 | 5 (55.5%) | 4 (45.5%) |
| CAPH | 21 | 8 (36.3%) | 13 (61.9%) |
| Cotrimoxazole | 21 | 19 (90.4%) | 2 (9.6%) |
| Ampicilline | 27 | 24 (88.8%) | 3 (11.2%) |
| Ciprofloxacin | 31 | 19 (61.2%) | 12 (38.8%) |
| **Nitrofurantoin** | **18** | **5 (27.8%)** | **13 (72.2%)** |
| Augmentin | 40 | 33 (82.5%) | 7 (17.5%) |
| Ceftriaxone | 29 | 23 (79.3%) | 6 (20.7%) |
| Ceftazidime | 23 | 15 (65.2%) | 8 (34.8%) |
| **Amikacin** | **16** | **0** | **16 (100%)** |
| **Meropenem** | **5** | **0** | **5 (100%)** |
| **Cefoxitine** | **3** | **0** | **3 (100%)** |
| **vancomycine** | **7** | **0** | **7 (100%)** |

**Table 3.**
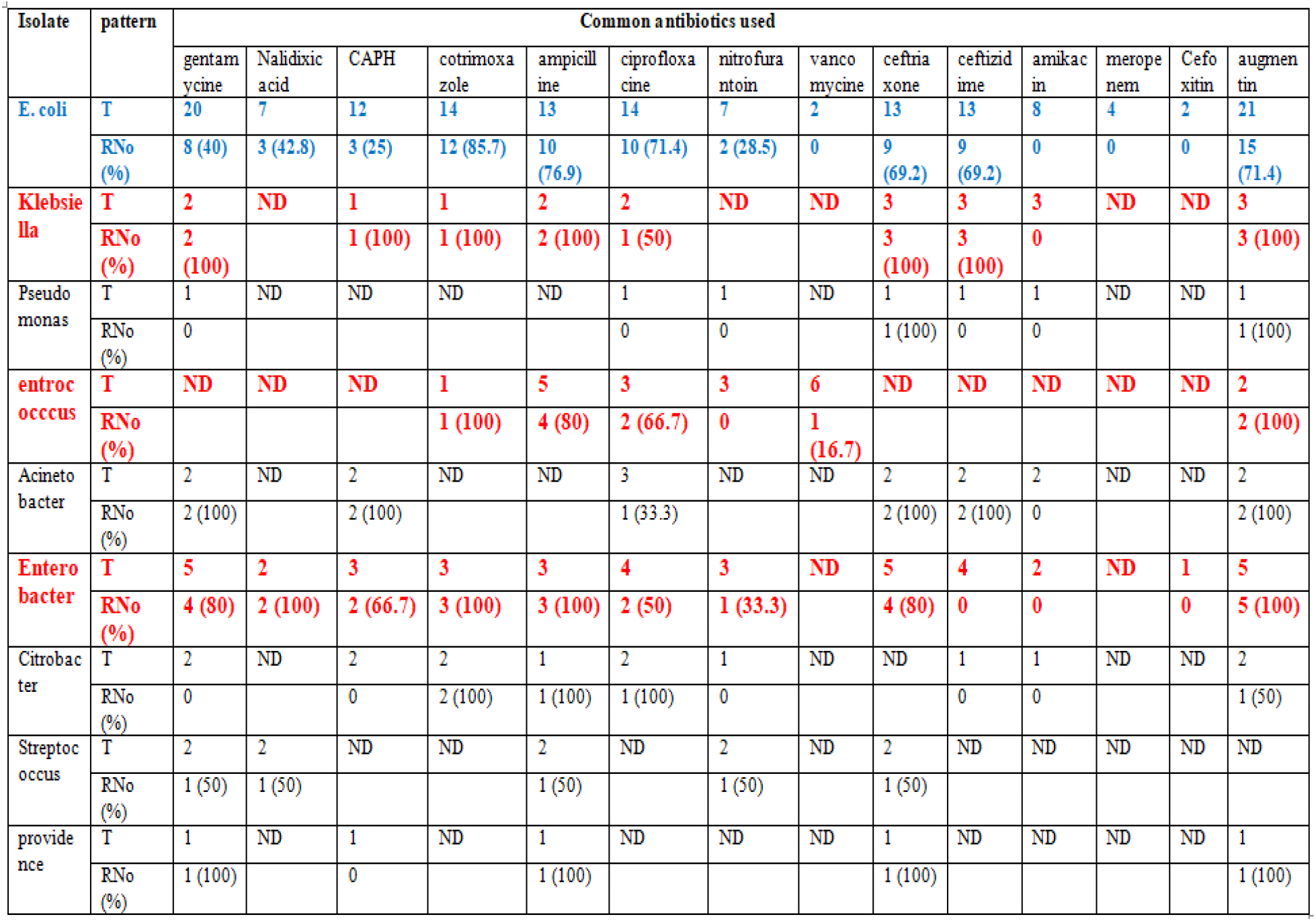
Overall proportions of the isolates’ resistant to all tasted antimicrobials, Tikur Anbessa specialized hospital emergency department from, Jan1 2015 to Dec 30, 2016, Addis Ababa Ethiopia ……

Majority (100%) of E. coli isolates were susceptible to Vancomycin, Meropenem, Amikacin, and Cefoxitin with no resistance rate. The other three common isolates were sensitive to Nitrofurantoin, Cefoxitin, Amikacin, and Vancomycin with resistance rates of 0% - 17%.

### 4. Multidrug resistance pattern of Uropathogens

In this study, the overall resistance pattern of isolated uropathogens to more than one antimicrobial was 86% and only 1(2 %) isolate was sensitive to all antimicrobials for which sensitivity was checked. The antimicrobial resistance rates of common isolates to more than one antibiotic were E. coli (80%), Enterococcus (83.3%), Enterobacter (100%), and klebsiella (100%) (Table4).

**Table4.**
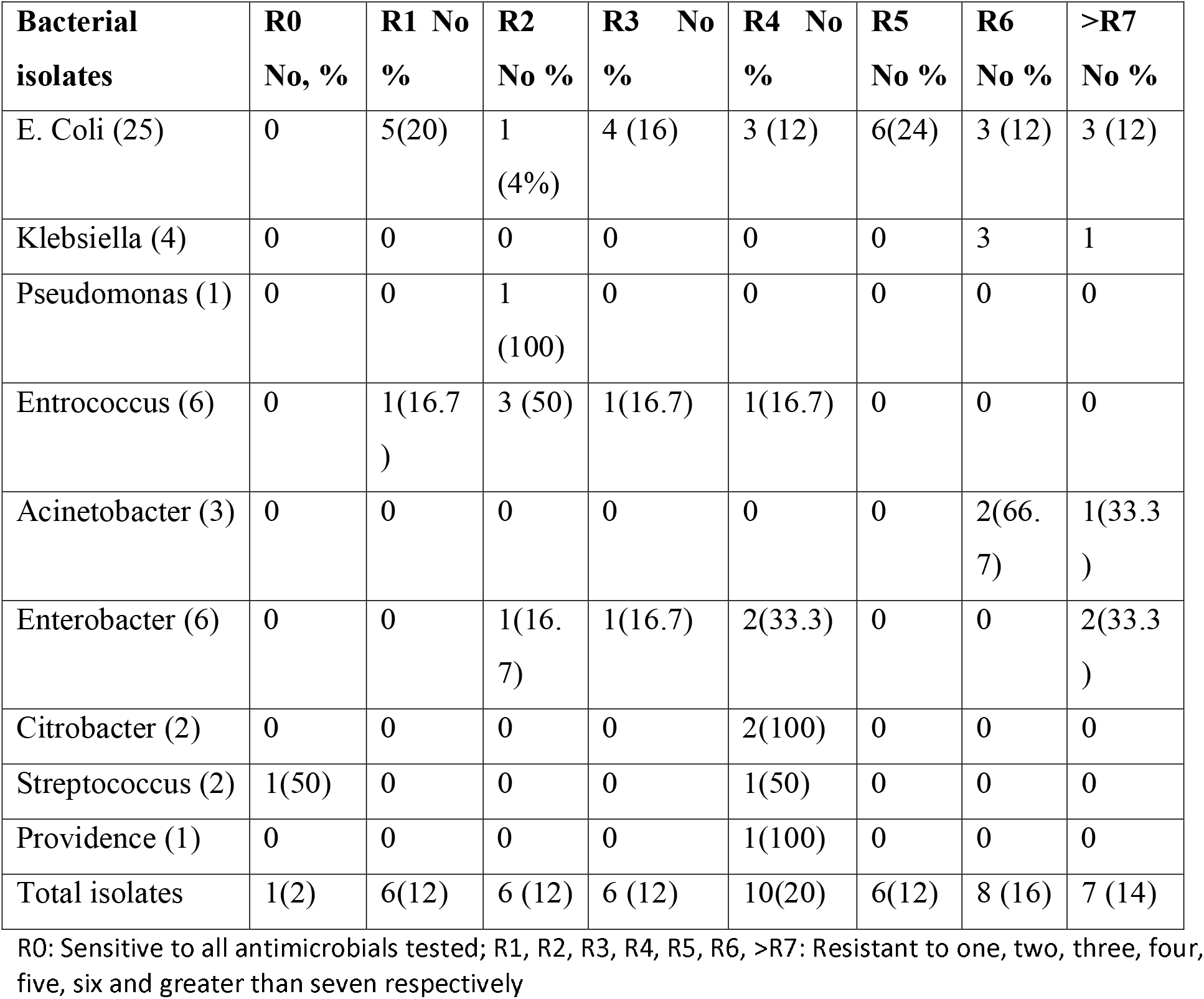
Multi Drug resistance pattern of isolates of urine culture, Tikur Anbessa Specialized Hospital emergency department from jan1 2015 to Dec 30 2016, Addis Ababa Ethiopia

## DISCUSSION

While urinary tract infections remain one of the most common disease patterns diagnosed worldwide, the management of these infections has become challenging due to the ever-rising emergence of antimicrobial drug resistance. The overall isolation rate of uropathogens in this study was found to be 22.7%, which is slightly lower than the rates reported from previous research conducted at this hospital [17]. However, the rate was similar to that of Adugna et al. [18]. In this study, E. Coli was the most predominant bacterium isolated from urine, followed by Enterobacter spp., Enterococcus spp., and Klebsiella spp. The isolation rates of E. Coli, which is the most common isolate, and other pathogens in this study, were comparable to the rates documented previously and also strongly agreed with similar research [8, 13, 16, 19].

Statistically, a significant difference was not observed between the two genders, though most pathogens were isolated from females (P = 0.34). Physiological and anatomical differences are accounted for by the differences between males and females. Females are more susceptible to infection than males, a phenomenon explained by their short urethra and its anatomical proximity to the anal orifice [3, 4].

This study showed that gram-negative bacteria were more frequent (84%) than gram-positive bacteria (16%). Similar findings have been reported by Tiruneh et al. from Gondar, Ethiopia, and other studies done elsewhere [16, 19, 20, 21, 22]. Bacterial uropathogenic isolates from patients with UTIs revealed the presence of extremely high levels of single and multiple antimicrobial resistance to commonly prescribed drugs.

The most frequently isolated bacterial isolates were found to be highly resistant to ciprofloxacin but sensitive to Nitrofurantoin. This result is different from the one documented previously in this hospital [17], in which the isolated organisms were highly sensitive to the aforementioned antimicrobials, even though sensitivity to Vancomycin, Meropenem, and Cefoxitin was not tested. Similar findings are reported by Gondar, including that bacterial isolates are highly resistant to CAPH [16, 12].

Enterococcus spp., Enterobacter spp., and Klebsiella spp. were found to be resistant to Ceftriaxone, Ceftazidime, Ciprofloxacin, Cotrimoxazole, Gentamicin, and Nalidixic Acid but sensitive to Nitrofurantoin, Vancomycin, and Cefoxitin. This finding is in contrast with a study done by Kibret et al. [1], in which both gram-negative and gram-positive isolates are very sensitive to Gentamicin and Ciprofloxacin. Additionally, reports from a previous study done in the same hospital [17], show that gram-negative isolates were mainly sensitive to Ciprofloxacin, Norfloxacin, and Nitrofurantoin, a finding that is also in contrast with ours. In this study, both gram-negative and gram-positive isolates were sensitive to Vancomycin at an equal rate, which is indicative of the gram-negative converse in the empirical use of Vancomycin.

In our study, antibiotic susceptibility data revealed that virtually all isolates (98%) were resistant to at least one antibiotic, and most of the isolates (86%) were resistant to two or more antibiotics. These findings were extremely higher than the resistance rates that were observed in other studies [16, 17], the reason for which might be the use of antibiotics and antimicrobial treatment prior to presentation to the hospital.

### Conclusion and recommendation

This study outlined that urinary tract infections are most commonly caused by Gram-negative bacteria predominantly in females, E. coli is the most prevalent isolate, and the multidrug resistance of urinary tract infections was significantly high in the largest referral hospital in Ethiopia, Black Lion, with almost all bacteria tested for more than one drug resistance except for E. coli. Co-trimoxazole, Ampicillin, Augmentin, and Ceftriaxone were found to be resistant against almost all bacterial isolates tested. Most bacterial isolates were found to be sensitive to Vancomycin, Chloramphenicol, Amikacin, Meropenem, Cefoxitin, and Nitrofurantoin, indicating that these medications could be used as the first line of emperical treatment for UTIs in emergency rooms. However, using antibiotics indiscriminately for patients with complicated UTIs may increase the resistance rate and also lead to treatment failure, hence the prescriptions should be revised following the culture and sensitivity results.

### Limitation

A limitation of this study can be understood in light of the following: Black Lion Hospital, as one of the largest tertiary units in the country, has a plethora of referral cases, the majority of which consist of debilitating comorbidities, like DM, malignancies, spinal cord injury, and septic shock, and not isolated UTO. This retrospective study is based on the results of routine microbiological tests carried out from 2015 through 2016, and so due to the nature of the retrospective analysis, tracing patients’ clinical settings was challenging and beyond the scope of the study. Additionally, the practice trends in the hospital are that culture and sensitivity tests are done for inpatients and not for those seeking outpatient care, which skews the caseload towards that of a complicated UTI.

## Data Availability

All data produced in the present work are contained in the manuscript

## Declarations

I declare that this thesis is entirely my own except where clearly stated otherwise, it has not been submitted in whole or in part in any previous research.

## Ethics consideration

Ethical approval for the study has been sought and obtained from Addis Ababa University Ethical Review board.

## Consent to publish

Not applicable

## Authors contribution

**Name:** Yared Boru, MD.

**Contribution:** This author helped with conceptualization, methodology, analysis, review and preparation of the manuscript.

**Name:** Dominick Shelton, MD.

**Contribution:** This author helped with conceptualization, methodology, review and editing of the manuscript.

**Name:** Finot Debebe, MD.

**Contribution:** This author helped with conceptualization, methodology, analysis, review and editing of the manuscript.

**Name:** Aklilu Azazh, MD.

**Contribution:** This author helped with conceptualization, supervision, review and editing of the manuscript.

**Name:** Fitsum Kifle, Msc.

**Contribution:** This author helped with methodology, review, editing and preparation of the manuscript.

**Name:** Hywet Engida, MD.

**Contribution:** This author helped with conceptualization, methodology, supervision and editing of the manuscript.

## Data availability

The article and its supporting files contain the datasets that support its conclusions. Any additional material can be obtained upon reasonable request.

## Appendices

